# Inhaled budesonide in the treatment of early COVID-19 illness: a randomised controlled trial

**DOI:** 10.1101/2021.02.04.21251134

**Authors:** Sanjay Ramakrishnan, Dan V. Nicolau, Beverly Langford, Mahdi Mahdi, Helen Jeffers, Christine Mwasuku, Karolina Krassowska, Robin Fox, Ian Binnian, Victoria Glover, Stephen Bright, Christopher Butler, Jennifer L Cane, Andreas Halner, Philippa C Matthews, Louise E Donnelly, Jodie L Simpson, Jonathan R Baker, Nabil T Fadai, Stefan Peterson, Thomas Bengtsson, Peter J Barnes, Richard EK Russell, Mona Bafadhel

**Author notes:** Senior and corresponding author, **Corresponding author**: Prof Mona Bafadhel, **Address**: Respiratory Medicine Unit, NDM Research Building, Nuffield Department of Medicine, University of Oxford, Old Road Campus, Roosevelt Drive, Oxford, OX3 7FZ, United Kingdom, **Email**. Joint first authors (Sanjay Ramakrishnan, Dan V. Nicolau).

## Abstract

**Background:** Multiple early hospital cohorts of coronavirus disease 2019 (COVID-19) showed that patients with chronic respiratory disease were significantly under-represented. We hypothesised that the widespread use of inhaled glucocorticoids was responsible for this finding and tested if inhaled glucorticoids would be an effective treatment for early COVID-19 illness.

**Methods:** We conducted a randomised, open label trial of inhaled budesonide, compared to usual care, in adults within 7 days of the onset of mild Covid-19 symptoms. The primary end point was COVID-19-related urgent care visit, emergency department assessment or hospitalisation. The trial was stopped early after independent statistical review concluded that study outcome would not change with further participant enrolment.

**Results:** 146 patients underwent randomisation. For the per protocol population (n=139), the primary outcome occurred in 10 participants and 1 participant in the usual care and budesonide arms respectively (difference in proportion 0.131, p=0.004). The number needed to treat with inhaled budesonide to reduce COVID-19 deterioration was 8. Clinical recovery was 1 day shorter in the budesonide arm compared to the usual care arm (median of 7 days versus 8 days respectively, logrank test p=0.007). Proportion of days with a fever and proportion of participants with at least 1 day of fever was lower in the budesonide arm. Fewer participants randomised to budesonide had persistent symptoms at day 14 and day 28 compared to participants receiving usual care.

**Conclusion:** Early administration of inhaled budesonide reduced the likelihood of needing urgent medical care and reduced time to recovery following early COVID-19 infection.

(Funded by Oxford NIHR Biomedical Research Centre and AstraZeneca; ClinicalTrials.gov number, NCT04416399)

**Research in context:** *Evidence before this study:* The majority of interventions studied for the COVID-19 pandemic are focused on hospitalised patients. Widely available and broadly relevant interventions for mild COVID-19 are urgently needed.

*Added value of this study:* In this open label randomised controlled trial, inhaled budesonide, when given to adults with early COVID-19 illness, reduces the likelihood of requiring urgent care, emergency department consultation or hospitalisation. There was also a quicker resolution of fever, a known poor prognostic marker in COVID-19 and a faster self-reported and questionnaire reported symptom resolution. There were fewer participants with persistent COVID-19 symptoms at 14 and 28 days after budesonide therapy compared to usual care.

*Implications of all the available evidence:* The STOIC trial potentially provides the first easily accessible effective intervention in early COVID-19. By assessing health care resource utilisation, the study provides an exciting option to help with the worldwide pressure on health care systems due to the COVID-19 pandemic. Data from this study also suggests a potentially effective treatment to prevent the long term morbidity from persistent COVID-19 symptoms.

## Introduction

COVID-19 is the most serious pandemic for over 100 years and with increasing mortality and morbidity worldwide. Other than age, obesity, and gender^1,2^, there are no clear predictors to forecast who will deteriorate needing hospital-based care. The onset of COVID-19 illness is almost always mild^3^ giving a potential window to intervene prior to the development of severe disease^1,2^.To date, almost all studies have focussed on investigating and treating severe and hospitalised COVID-19 infection^4^. However, there is currently little knowledge on therapeutic targets in early COVID-19 infection to prevent progression and clinical deterioration.

In early reports describing COVID-19 infection from China^1,2^, Italy^5^ and the United States^6^, there was a significant under representation of patients with asthma and chronic obstructive pulmonary disease (COPD) in patients hospitalised with COVID-19. We hypothesized that this may be due to the widespread use of inhaled glucocorticoids in these patients^7^. Furthermore, the main indication for the use of inhaled glucocorticoids in patients with asthma and COPD is to reduce exacerbations, which are recognised to be often viral in etiology^8,9^. *In-vitro* studies have shown that inhaled glucocorticoids reduce the replication of SARS-CoV-2 in airway epithelial cells^10^; in addition to the downregulation of expression of angiotensin converting enzyme-2 (ACE2) and transmembrane protease serine 2 (TMPRSS2) genes which are critical for viral cell entry^11^.

Here we present the analysis of the Steroids in COVID-19 (STOIC) trial, a phase 2 trial designed to evaluate the efficacy of inhaled budesonide in individuals with early COVID-19 illness in the community. We examined the effect of inhaled budesonide on likelihood of urgent care or hospitalisation, clinical recovery, temperature and oxygenation. We also evaluated the effect of inhaled budesonide on SARS-CoV-2 viral load.

## Methods

### Trial Design

The STOIC trial is a randomised, open label, parallel group phase 2 clinical trial conducted in the community in Oxfordshire, United Kingdom. Adults over the age of 18, with symptoms suggestive of COVID-19 within 7 days were eligible for inclusion. Participants were excluded if they had a known allergy or contraindication to the interventional medicinal product, or if they had recent use (within 7days) of inhaled or systemic glucocorticoids. Recruitment for the study was via local primary care networks, local COVID-19 testing sites and via multi-channel advertising. Volunteers were able to contact the study staff using the advertised phone numbers or via e-mail and all participant information was publicly available on the study website (www.stoic.ndm.ox.ac.uk). The trial was registered on clinicaltrials.gov (NCT04416399).

### Study intervention and assessments

Participants who met the inclusion criteria were randomised to usual care (UC) or intervention with budesonide (BUD) dry powder inhaler (Pulmicort Turbuhaler, AstraZeneca) at a dose of 800µg twice a day. Participants were seen at their homes at randomisation (day 0), day 7 and day 14 by a trained respiratory research nurse to obtain written informed consent, provide inhalers and to obtain nasopharyngeal swabs for SARS-CoV-2 reverse transcription polymerase chain reaction (RT-PCR) testing (see **supplementary methods** for further details). Each participant received a paper symptom diary, calibrated pulse oximeter and thermometer for daily home monitoring. All participants were telephone contacted daily to record oxygen saturations, temperature and assessed for any adverse events by the study team. Participants allocated BUD were asked to stop taking the inhaler when they felt they had recovered (self-reported symptom recovery) or if they hit primary outcome; and all participants ceased daily monitoring (including daily telephone calls) when symptoms had recovered (again self-reported symptom recovery) or if the primary outcome was achieved. Finally, at day 28, all study participants were seen in the trial centre and serum SARS-CoV-2 antibody testing was performed.

### Outcomes

The primary outcome was defined as a COVID-19-related Urgent Care visit, Emergency Department assessment or hospitalisation. During the pandemic, members of the public in the United Kingdom, were encouraged to contact a government telephone advice line prior to attending the emergency department and COVID-19 specific general practice hubs were created for patients who were deteriorating at home, for medical treatment including transfer to hospital. Secondary outcomes included clinical recovery as defined by self-reported time to symptom resolution; viral symptoms measured by the Common Cold Questionnaire (CCQ)^12^ and the InFLUenza Patient-Reported Outcome (FluPRO®)^13^ questionnaire; blood oxygen saturations and body temperature; and SARS-CoV-2 viral load.

### Randomisation and Statistical Analysis

The statistical packages R (**R** Core Team (2020). **R**: A language and environment for statistical computing. **R** Foundation for Statistical Computing, Vienna, Austria. URL https://www.R-project.org/), Gauss ((https://www.aptech.com/), and SAS v9.4 (https://www.sas.com/en_us/home.html) are used. Descriptive statistics are used to describe variables between the groups in the BUD arm and the UC arm. Appropriate parametric or non-parametric statistical tests are performed. For continuous variables, the difference between treatments in the means or medians and the corresponding 95% confidence interval are reported. For continuous variables, ANCOVA models (t-tests) adjusted for stratification factors or Wilcoxon rank sum tests are applied to compare the BUD and UC arm. For categorical variables the number (and percentage) of patients in each category are reported for each treatment group and chi-squared tests will be used for comparing treatment arms. Time to self-reported clinical recovery and FLUPro® symptom recovery are analysed using the Kaplan Meier method and presented as the median time to event. Comparison is performed with a log rank test where participants with primary outcome are censored at Day 28 when not meeting the event. Participants are randomly allocated to UC or BUD, stratified for participant age,(≤40 years/ >40 years) gender, and number of co-morbidities (≤1/ ≥2). The randomisation sequence was created using a random number generation function and allocation to each arm was performed through block randomisation in a 1:1 ratio. All tests are performed at a 5% 2-sided significance level and all comparative outcomes are presented as summary statistics with 95% confidence intervals and reported in accordance with the CONSORT Statement. Missing data from study visits and daily monitoring are handled by last observation carried forward (LOCF) for temperature, oxygen saturations and time to FLUPro® symptom resolution. For FLUPro® total score and individual domain time-series plots, missing data was handled by imputation of zero score when self-reported symptom resolution occurred or LOCF when this was not available. Stochastic simulations of a “virtual” trial with the same study design, primary endpoint and duration, and community detection are presented in full in the **supplementary materials**. P-values are reported to a minimum of 3 decimal places. Further full details are available in the **supplementary material**.

### Sample size estimation

At study inception in March 2020 and using published data available at the time^1,2^, we assumed that 20% of all COVID-19 illness is severe and would require hospitalisation. Using 80% power at 0.05 level, we required 199 patients in each arm to demonstrate a 50% reduction of urgent care visits or hospitalisations. The primary outcome was analysed for both the per-protocol (PP) and intention to treat (ITT) population. The PP population is defined as the population that received the study treatment and had at least 1 day of study observations; the ITT population is defined as all participants that were randomised to a study arm.

### Institutional Review

The trial was sponsored by the University of Oxford, and was approved by the Fulham London Research Ethics Committee (20/HRA/2531) and the National Health Research Authority. All participants provided written informed consent. The study team requested an independent statistical monitoring committee review on the 9^th^ of December 2020 due to reduced recruitment after the second national lockdown in England; implementation of the COVID-19 vaccine; and ethical consideration of the primary outcome. *A priori* stop criteria were used to determine futility of further recruitment (**see statistical analysis plan**).

## Results

### Participants

From July to December 2020, 146 participants were randomised, of which 139 were in the PP analysis, with 70 and 69 in the BUD and UC arm respectively (**see figure 1)**. Participant characteristics were similar between the study arms, as shown in **table 1** (see **supplementary table 1 for ITT population**). COVID-19 infection occurred in 94% measured by RT-PCR. The median duration of symptoms prior to randomisation was three days. The median time to symptom resolution was eight days. Inhaled budesonide was taken for a median (IQR) duration of 7 (4 to 10.5) days.

**Table 1.** Demographics and clinical characteristics of study participants in the per-protocol population

| Characteristic | Budesonide (n = 70) | Usual care (n = 69) |
| --- | --- | --- |
| Age, years | 44 (19-71) | 46 (19-79) |
| Female sex, no. (%) | 39 (56%) | 41 (59%) |
| White ethnicity, no. (%) | 65 (93%) | 64 (93%) |
| Body mass index, kg/m<sup>2</sup> | 27 (19-38) | 26 (18-42) |
| Number of co-morbidities no.# | 1 (0-4) | 1 (0-4) |
| Duration of symptoms, days# | 3 (1-7) | 3 (0-7) |
| Evidence of COVID positive status, no. (%) | 66 (94) | 65 (94) |
| Presence of symptoms at baseline, no. (%) |  |  |
| Cough | 55 (79%) | 48 (70%) |
| Fever | 49 (70%) | 44 (64%) |
| Headache | 40 (57%) | 38 (55%) |
| Fatigue | 32 (46%) | 23 (33%) |
| Loss of sense of smell/taste | 25 (36%) | 30 (43%) |
| Gastrointestinal symptoms | 11 (16%) | 12 (17%) |
| Breathlessness | 11 (16%) | 11 (16%) |
| Myalgia | 6 (9%) | 10 (14%) |
| Nasal symptoms | 3 (4%) | 5 (7%) |
| Sore throat | 0 (0%) | 2 (3%) |
| Chest pain/tightness | 4 (6%) | 1 (1%) |
| Other | 7 (10%) | 8 (12%) |
| Highest temperature recorded*# | 36.6 (35.2 to 39.0) | 36.6 (35.5- 38.3) |
| Lowest Oxygenation recorded as % saturation*# | 96 (84-99) | 96 (90-99) |
| SARS-CoV-2 viral cycle threshold¥ | 32.6 (22.4-39.4) | 31.8 (15.6-40.0) |

Data presented as mean (SD) or mean (range) unless otherwise stated; \*at randomisation; ¥median (IQR); #median (range); \$ mean (95%CI)

**Figure 1.**
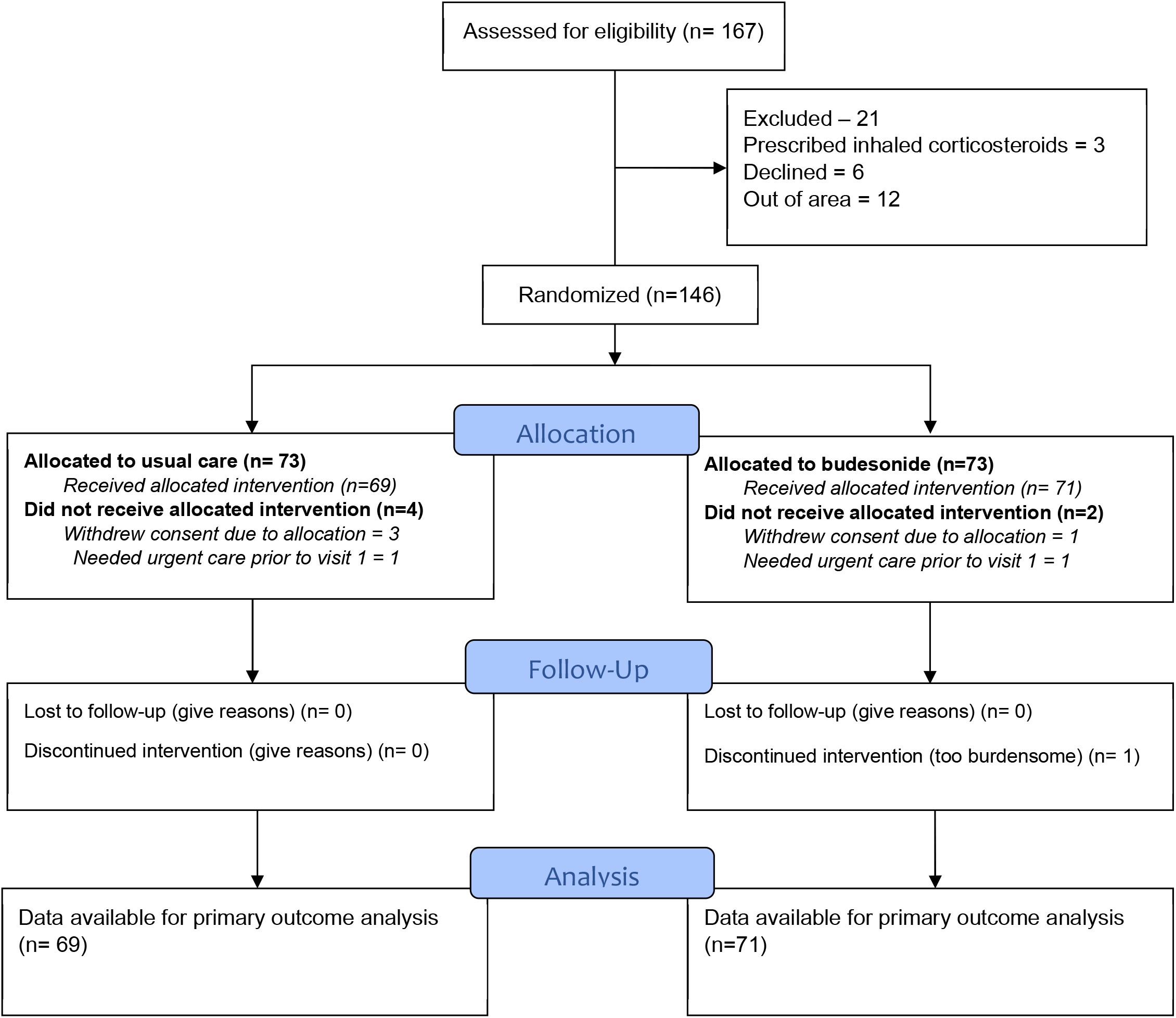
Consort flow diagram

### Conditional power

Simulations using bootstrap was performed to determine the conditional power for an evaluation of an early stop, using the *a priori* decisions described in the supplement at approximately 146 patients. Estimated power was >99% using both the total population (N=124) and, at the time of the simulation, the known subgroup of COVID-19 positive patients (N=78).

### Primary outcome

In the PP analysis, the primary outcome occurred in 10 participants in the UC arm versus 1 participant in BUD arm (difference in proportions 0.131, 95% CI (0.043, 0.218), p=0.004) indicating a relative risk reduction of 90% for BUD. The number needed to treat with inhaled budesonide to reduce COVID-19 related urgent care/hospitalisation was 8. Sensitivity analysis in participants with confirmed COVID-19 showed that the difference in proportions was 0.125, 95% CI (0.035, 0.216), p=0.007. There was no difference in participants with a primary outcome event compared to those without (**see supplementary table 2**). For the ITT population, the primary outcome occurred in 11 participants in the UC arm and 2 participants in the BUD arm (difference in proportion 0.123, p=0.009).

### Secondary outcomes

Self-reported clinical recovery was 1 day quicker with BUD compared to UC (median of 7 days versus 8 days, logrank test p=0.007, **see figure 2**). The mean time to recovery, in days was 8 and 11 in the BUD and UC arm respectively. Further sensitivity analysis for clinical recovery in participants with confirmed COVID-19 status showed a similar median times to recovery (7 vs. 8 days, p=0.039) **(see supplementary figure 1)**. At day 14, self-reported symptoms were present in 10% (n=7) of participants randomised to BUD compared to 30% (n=21) of participants randomised to UC (difference in proportion 0.204, p=0.003).

**Figure 2.**
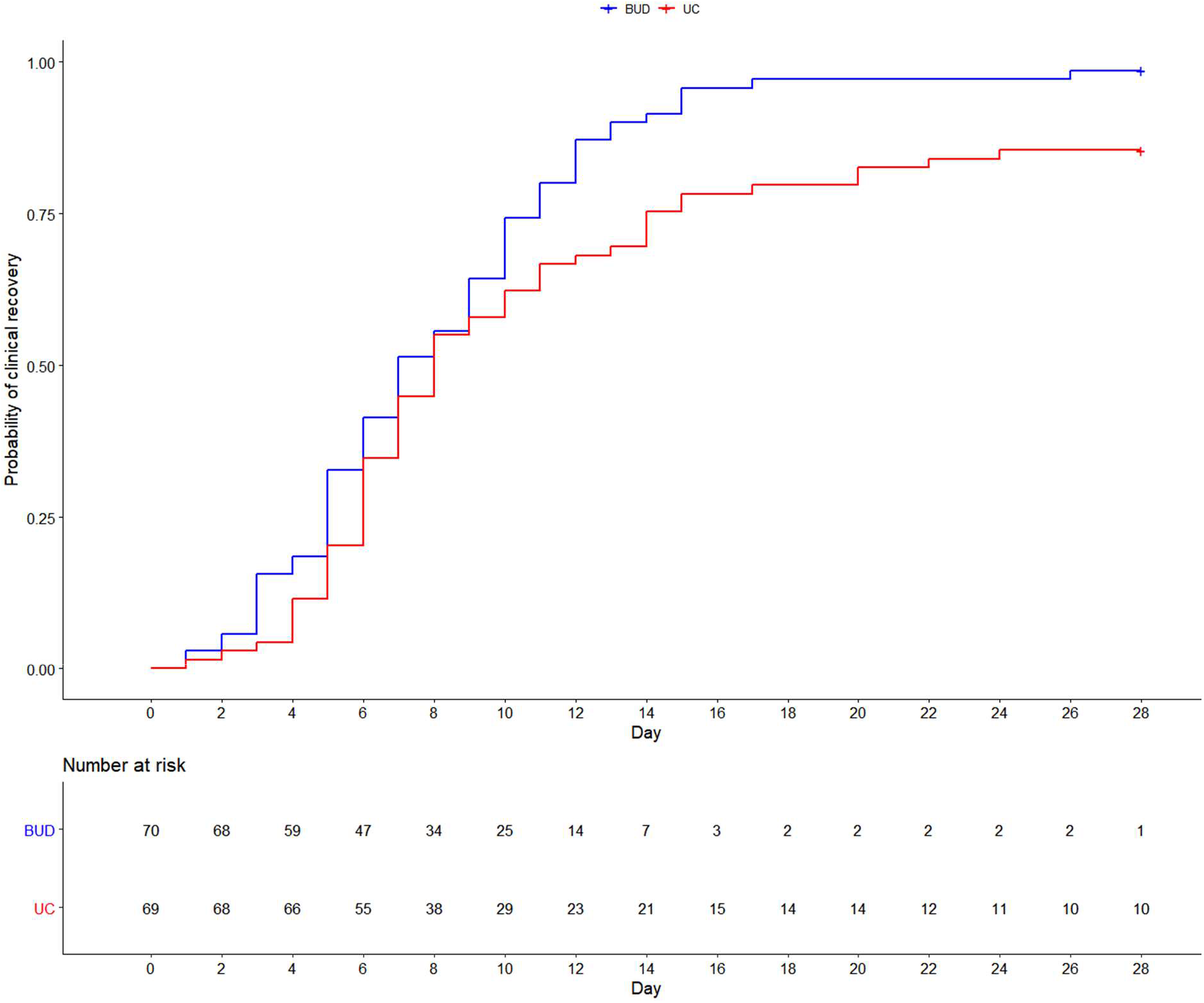
Time to self-reported clinical recovery of per protocol population using data censoring for primary outcome. BUD = budesonide; UC = usual care

The mean proportion of days with a documented fever (≥37.5 C) during the first 14 days, was 2.1% in the BUD and 7.7% in the UC arms (Wilcoxon test, p= 0.051). The percentage of participants with at least 1 day of fever was 11% and 23% in the BUD and UC arm respectively (Fishers test, p=0.076). Violin plots, showing distribution of pooled highest temperatures are presented in **figure 3**, with a statistically higher mean in the UC arm (mean difference 0.49, 95%CI 0.32 to 0.66, p<0.001). Temperature plots relative to the day of randomisation showed that temperature fell quicker in the BUD compared to UC arm **(see supplementary figure 2)**. The median (IQR) proportion of total days that participants required as needed anti-pyretics (paracetamol, aspirin, ibuprofen) in the BUD and UC arms was 27% (0 to 50) and 50% (15 to 71) respectively (Mann-Whitney test, p=0.025).

**Figure 3.**
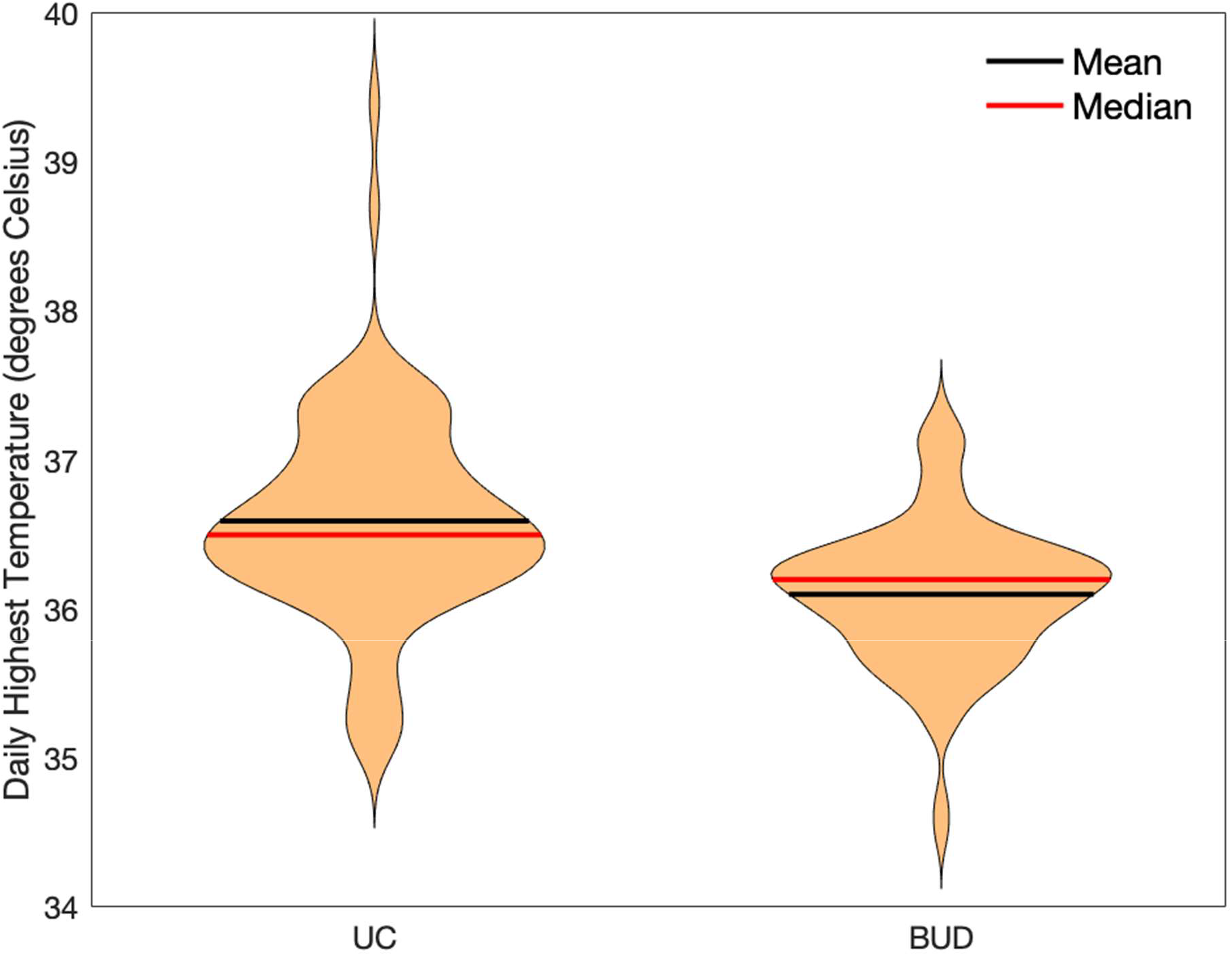
Violin plots illustrating pooled peak (maximum) temperature in participants in BUD and UC arms, with statistically significant difference in the mean temperature (mean difference 0.49, 95%CI 0.32-0.66, p<0.001). BUD = budesonide; UC = usual care

Symptom resolution at day 14, as defined by the FLUPro® user manual, occurred in 82% and 72% of the BUD and UC arms respectively (p=0.166); whilst the median time to symptom resolution as measured by the FLUPro® was 3 and 4 days in the BUD and UC arms respectively (logrank test p=0.080, **see supplementary figure 3**). The mean change (95%CI) in FLUPro® total score between day 0 and day 14 in the BUD and UC are -0.65 (−0.80 to -0.50) and -0,54 (−0.69 to -0.40) respectively (mean difference of -0.10, 95%CI of the difference -0.21 to -0.00, p=0.044). **Figure 4 (panels a-g**) shows the mean daily FLUPro® scores for the total symptom burden and individual domains. The mean change of the FLUPro® domains showed that systemic symptoms were significantly greater in BUD compared to UC (**supplementary table 3)**. The mean change in CCQ total score between day 0 and day 14 in the BUD and UC was -0.49 (−0.63 to -0.35) and -0.37 (−0.51, -0.24) respectively (mean difference of -0.12 (−0.21 to -0.02), p=0.016). The common cold questionnaire (CCQ) symptom daily mean score is presented as **supplementary figure 4**.

**Figure 4.**
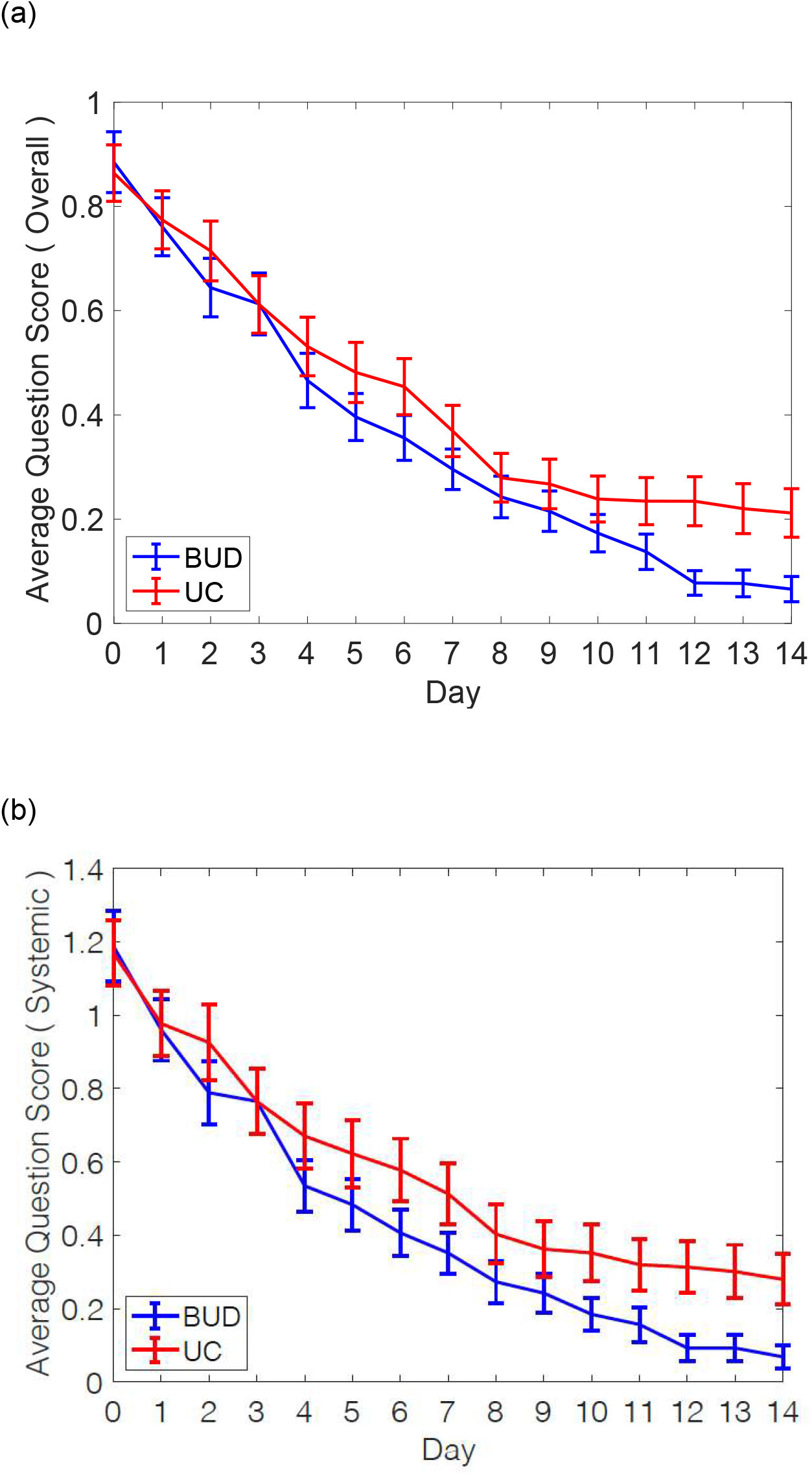
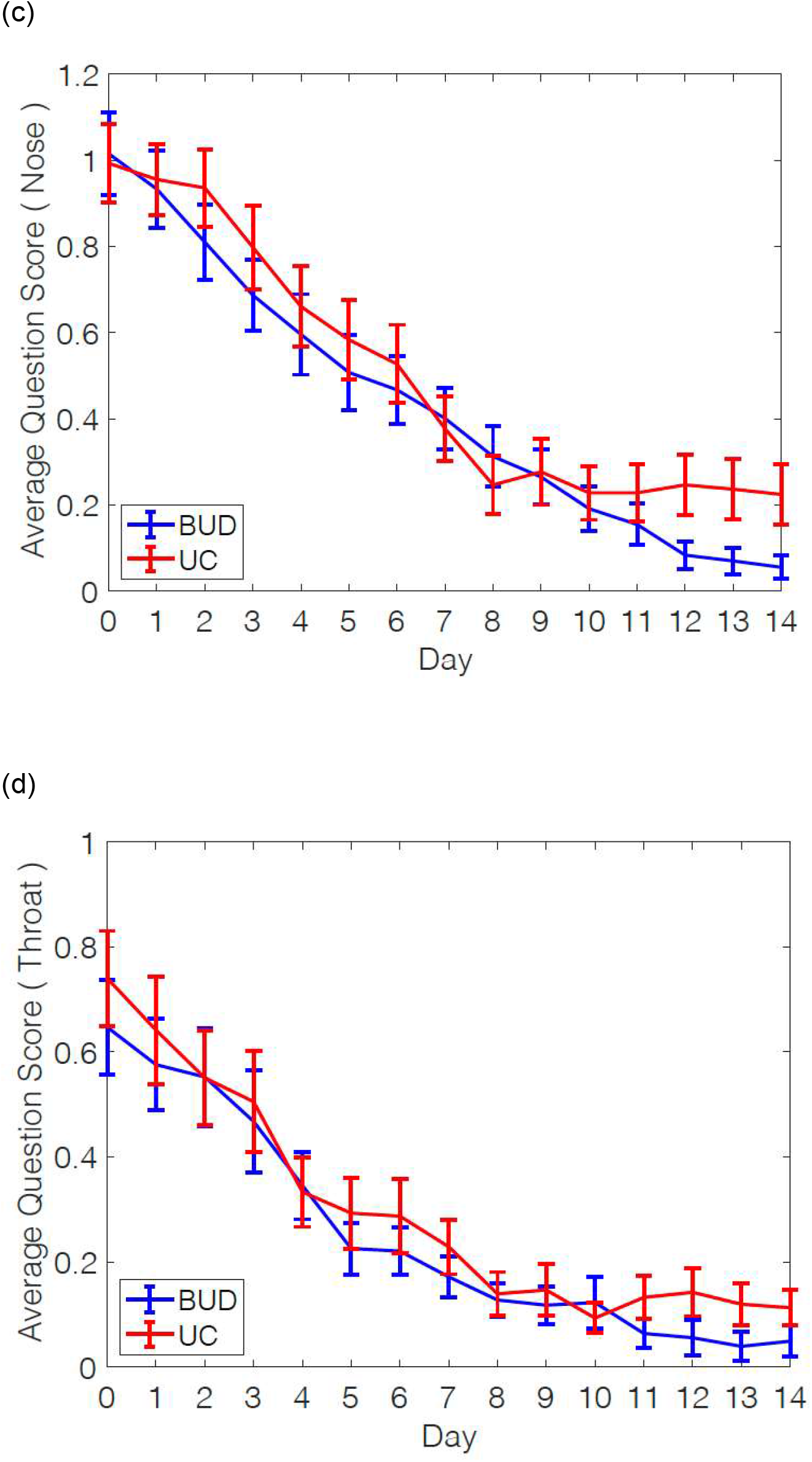
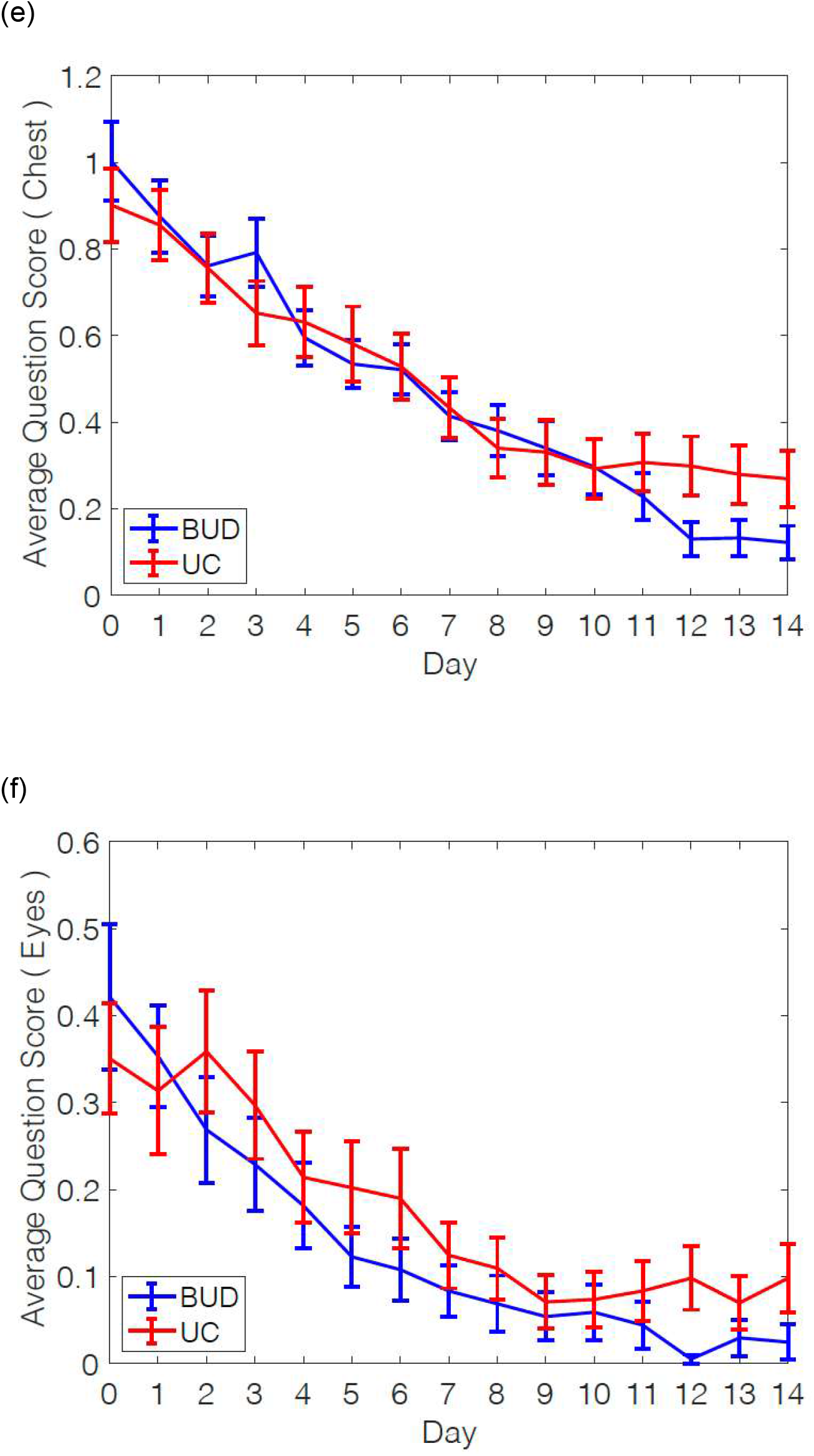
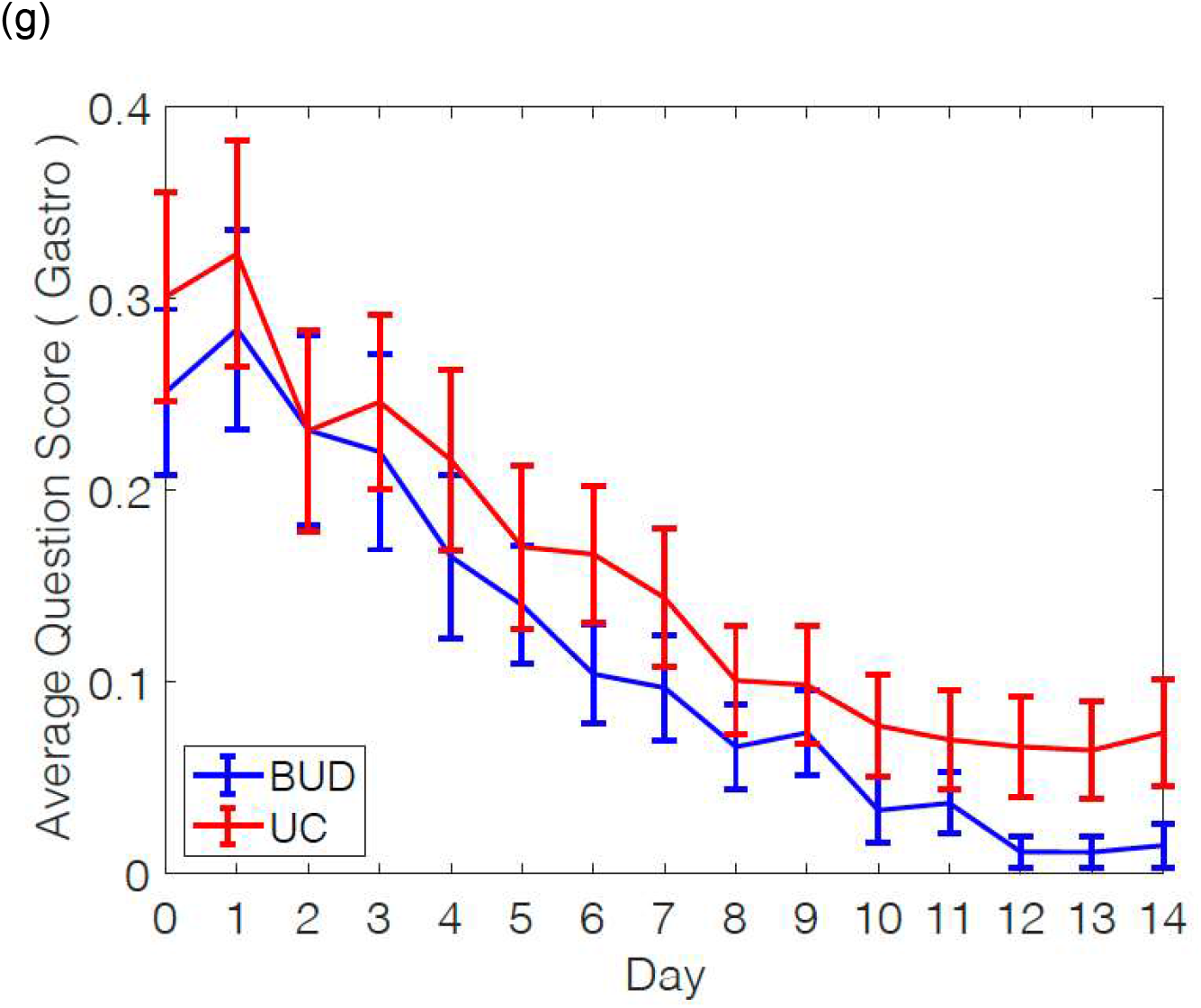
Daily mean scores over 14 days using the FLUPro® questionnaire. Panel (a) total symptoms; (b) systemic symtoms (c) nasal symptoms; (d) throat; (e) chest; (f) eyes; (g) gastrointestinal. BUD = budesonide; UC = usual care.

The proportion of days with oxygen saturations ≤94%, during the first 14 days, was 19% and 22% in the BUD and UC arm respectively (Wilcoxon test p=0.627). During the first 14 days 59% and 58% in the BUD and UC arms had at least one day with oxygen saturations ≤94%.

The median (IQR) cycle threshold (CT) nasopharyngeal SARS-CoV-2 viral load at Day 0, day 7 and day 14 was 32.1 (21.7-40.0), 35.25 (32.4-40.0) and 36.4 (34.2-40.0) respectively. There was a significant difference in CT reduction between visit 1 and 2 for both study arms (Wilcoxon matched pairs P<0.001, **see supplementary figure 5**); but no difference in reduction between groups (mixed effect ANCOVA p=0.354).

**Figure 5.**
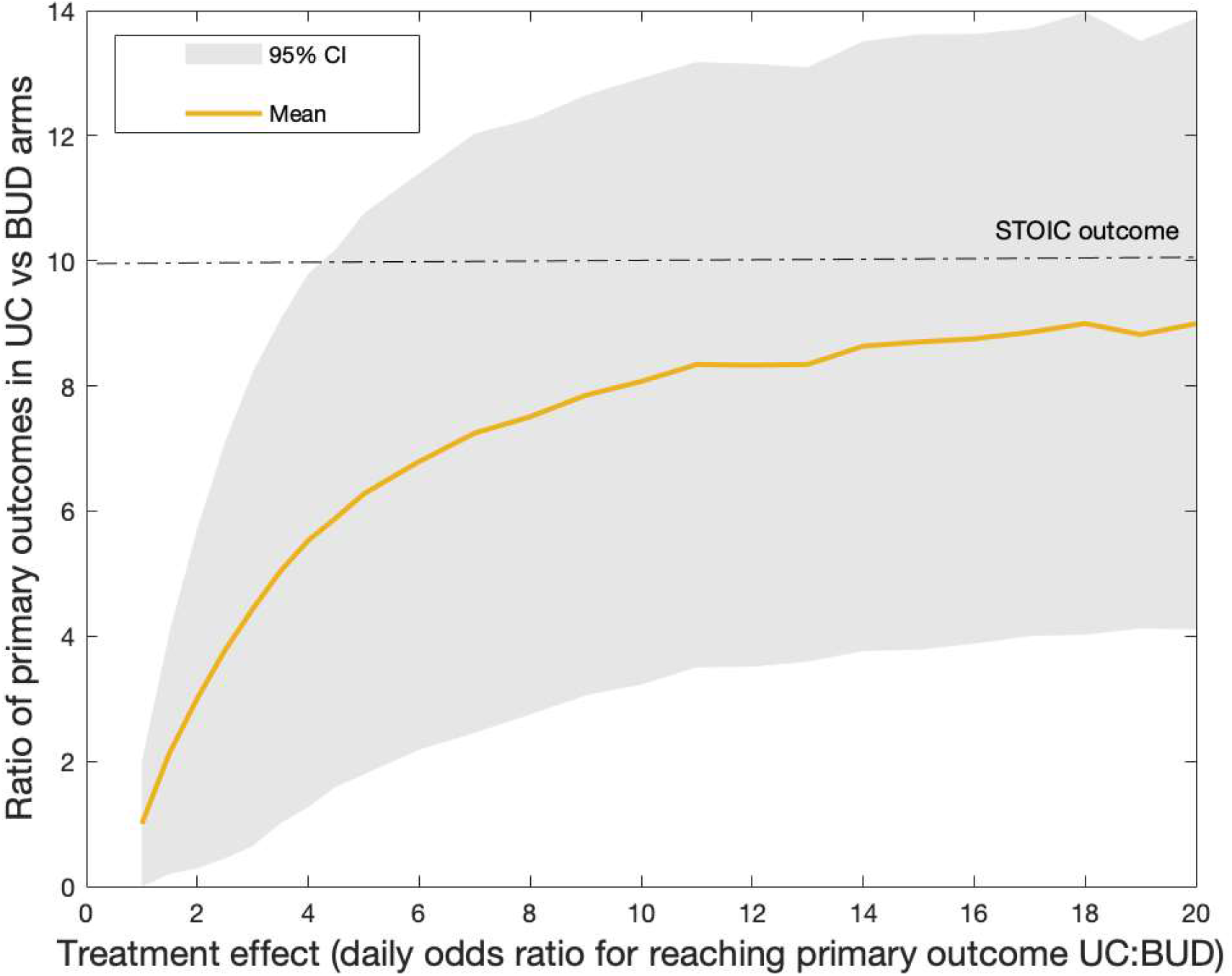
Relationship between treatment effect, here defined as the daily ratio of the odds of reaching primary outcome (PO) in the UC vs BUD arms (horizontal axis) and the ratio of primary event outcomes in the UC vs BUD arms at the completion of the trial (vertical axis). Plots derived from numerical simulations of the stochastic “virtual twin” trial. These results indicate that in order to observe our findings (shown by the dotted line), the daily treatment effect needed represents approximately 3000% (30x) reduction in the daily odds of reaching primary outcome.

### Further analysis

Stochastic simulations, in a ‘virtual twin’ study design, demonstrated that the daily odds ratio of reaching the primary outcome, with BUD reduced by 3000% (**see figure 5** and **supplementary materials)**.

## Discussion

We have demonstrated that the inhaled glucocorticoid, budesonide, given for a short duration, may be an effective treatment of early COVID-19 disease in adults. This effect, with a relative reduction of 90% of clinical deterioration is equivalent to the efficacy seen following the use of COVID-19 vaccines^14,15^ and greater than that reported in any treatments used in hospitalised and severe COVID-19 patients^16-18^. The broad inclusion criteria make this study intervention relevant to health care systems worldwide. Inhaled budesonide is a simple, safe, well studied, inexpensive and widely available treatment. The number of participants needed to treat to prevent increased health care resource utilization is 8, and combined with the short treatment period required to achieve benefit, makes this potentially an affordable and scalable intervention for early COVID-19. This is especially significant in low- and middle-income countries where the majority of currently approved COVID-19 treatments are unlikely to ever reach patients as a consequence of variable healthcare systems^19^. For example, although dexamethasone is a widely available and cheap medicine, and is effective in reducing mortality in severe and intensive care related COVID-19, this is unfortunately largely irrelevant in countries which have limited intensive care or hospital capacity^20,21^. Furthermore, in high income countries, inhaled budesonide could work as an adjunct to reduce pressure on health care systems until widespread SARS-CoV-2 vaccination can be achieved. Unlike with vaccines, the efficacy of inhaled budesonide is unlikely to be affected by any emergent SARS-CoV-2 variant^22^.

We selected this treatment intervention due to the unexpected observation of an under-representation of patients with asthma and COPD in hospitalised cohorts of COVID-19^23^. This finding from early hospitalised cohorts in Wuhan^1,2^ was at odds with prior respiratory viral pandemics, such as H1N1 influenza^24^. The common therapy between these lung diseases is inhaled glucocorticoids, either as a mono-, dual- or triple-constituent. Furthermore, inhaled glucocorticoids are among the most prescribed medicine of any class around the world, listed by the World Health Organisation (WHO) of essential medicines^25^. Moreover, evidence of the utility of inhaled glucocorticoids, in reducing viral exacerbations of asthma have been known for many decades^9^, whilst specifically, inhaled budesonide has shown effect at reducing rhinovirus replication *in-vitro*^26^. The efficacy of Dexamethasone in RECOVERY^17^, for severe disease also supports our findings, whilst there is plausibility that the immune-modulatory effect of inhaled glucocorticoids^27^, may also apply to any future viral epidemics. In our study, we found that inhaled budesonide also demonstrated benefit in the secondary outcomes, with quicker symptom resolution in those treated with budesonide either measured using a self-report of symptom recovery, or defined as normalisation of prospectively collected symptom scores measured using the FluPRO®^13^ or the CCQ^12^. The positive impact on temperature when used to treat early COVID-19 is further evidence that inhaled budesonide is modifying the disease process. Of note, there was a significantly greater population of participants randomised to budesonide who were free of symptoms at 28 days compared to participants randomized to usual care. In the United Kingdom, up to 20% of patients^28^ report persistent symptoms 5 weeks after COVID-19. This interesting finding, also suggests that intervention with an inhaled glucocorticoids might impact on rate of the persistent long-term symptoms in COVID-19 (“long COVID”); and should be investigated further in view of the significant long-term health and economic impact of long COVID.

There are limitations to our study. This is an open-labelled study, which was stopped early due to the impact of the national pandemic control measures and national prioritisation rules for clinical research trials in the UK. Our power calculations were made from the best available predictions in early 2020. Therapeutic randomised clinical trial design and sample size calculations are often dictated by statistical assumptions with treatment effect estimations based on the evidence of best available care. However, in trial design for a new disease, with no known effective treatment, statistical assumptions are thus arbitrary. In our study the event rate in the control arm was half of what was predicted, and this is consistent with subsequent primary care clinical trials performed in COVID-19^29-31^. We found that the budesonide treatment effect size, was larger than predicted; and independent statistical assessment concluded that the current sample size and treatment effect had a 99% power to reject the null hypothesis. Furthermore, the positive concordance of temperature and symptoms as secondary outcomes gives us confidence in our results. Our study design involved randomisation at home, with home visits for study assessments and a daily contact until symptom resolution by the study team. To our knowledge, the STOIC trial is first to assess daily physiological measures in early COVID-19. The robust study design meant very good participant retention and completion of symptom diaries. Stopping a study early is unusual and is a decision which is not taken without due diligence^32^ However, we ensured that *a priori* stop decision analysis was performed by an independent statistical team for statistical rigor. During these unprecedented times, our findings favouring budesonide, despite these limitations can not be ignored and requires urgent dissemination and validation; especially in the setting of a treatment that is relatively safe.

In conclusion, budesonide, a safe inhaled glucorticoid, appears to be an effective treatment for early COVID-19 infection which could be applicable to global healthcare systems.

## Data Availability

De-identified individual participant data and a data dictionary defining each field in the set, can be made available to others upon approval of a written request to the corresponding author. The request will be evaluated by a committee formed by a subset of co-authors to determine the research value. A data sharing agreement will be needed.

## Acknowledgements

We are grteful for the support of Rhys Painton, Sophie Morgan, Nick Thomas, Jennifer Quint, Philip Maini, Helen Byrne, Sunil Lakhani, Ruairidh Battleday and Adriaan Bax. We would also llke to thank Investec PLC for generously supporting the trial with personal protective equipment during times of supply shortages. We thank the Department of Health and Social Care who were instrumental in supporting the promotion of STOIC at Oxfordshire testing sites and the Oxford Respiratory Trials Unit.

## Author contributions

SR, BL, HJ, CM, KK, MM, and MB performed all study assessments, study visits and completed data entry.

DN, NTF, TB and SP provided statistical support.

TB and SP acted as independent statisticians.

MM and JB performed all PCR tests.

All authors contributed to the manuscript.

SR and MB vouch for all the data, analysis and submitted the final version of the manuscript for peer review.

