## Supplemental materials for "Inhaled budesonide in the treatment of early COVID-19 illness: a randomised controlled trial"

**Corresponding author:** Prof Mona Bafadhel

### Methods

***SARS-CoV-2 detection from Nasopharyngeal swabs:*** Nasopharyngeal swabs were collected from all participants at randomisation, day 7 and day 14. Participants were asked to perform self-swabs, using one swab collecting sample from the nasopharynx and throat. Research nurses performed the nasopharyngeal swab if the participant was unable to. Swabs were stored upon collection immediately in RNAse solution, for SARS-CoV-2 deactivation and transferred in protected collection pots to the main Respiratory Medicine laboratory at the University of Oxford.

***Virus extraction and quantification***: RNA was extracted from clinical samples using the QIAamp Viral RNA Mini Kit (Catalogue: 52906, Qiagen) following manufacturer's instructions. 420 µl of sample was extracted and eluted into 40 µl Buffer AVE. 10 µl of eluted RNA was assayed using the Taqman fast virus 1-step master mix (ThermoFischer Scientific, Loughborough, UK), utilising oligonucleotide primers (600 nM forward and 800 nM reverse per reaction) and fluorescent conjugated probes (two probes 100 nM each) (see supplementary table 1 for sequences) (Eurofins Genomics, Wolverhampton, UK) for the detection of the viral RNase P gene (RdRP) gene region of SARS-CoV-19 using the ABI 7500 SDS Instruments (Applied Biosystems). Virologic testing for SARS-CoV-2 infection was performed by quantitative real-time RT-PCR (RT-qPCR). Assay data are presented in cycle threshold (units). The limit of detection was 40 cycles.

*Sequences for primer and probes for RT-qPCR*

| |
|---|
| Dual Labelled probe P2: CAG GTG GAA CCT CAT CAG GAG ATG C |
| Dual labelled probe P1: CCA GGT GGW ACR TCA TCM GGT GAT GC |
| PCR primer RDRP F: GTG ARA TGG TCA TGT GTG GCG G |
| PCR primer RDRP R: CAR ATG TTA AAS ACA CTA TTA GCA TA |

***SARS-CoV-2 antibody detection:*** All participant samples were collected in 5ml serum separator tubes manufactured by BD Vacutainer®. Samples were labelled using a pseudo anonymised code and sent to the Oxford University Hospital NHS Foundation Trust microbiology laboratory within 6 hours of sample collection. Serology for IgG to nucleocapsid protein was performed using the Abbott Architect i2000 chemiluminescent microparticle immunoassay (Abbott, Maidenhead, UK). Antibody levels ≥1.40 arbitrary units were considered positive.

### Statistical Analysis

See attached Statistical analysis plan for primary and secondary analyses.

***Stochastic Simulations of a Virtual Twin of the STOIC Study:*** To further shed light on the data generated in the STOIC trial, we performed stochastic simulations of a "virtual" trial with the same design, primary endpoints and duration as STOIC. In our simulations, each of a number of virtual "patients" are recruited (R) assigned to either the BUD or UC arms, as illustrated in illustration 1. The simulation visits each virtual patient once per simulation day. The patient may stay in the BUD or UC "state" or transition to drop-out (DO), recovery (RES) or reaching primary end-point (PO), with probabilities for each transition pre-set, as illustrated (and summing to 1). A random number between 0 and 1 will be generated on each such day and the decision which transition to make is made in accordance to its value.

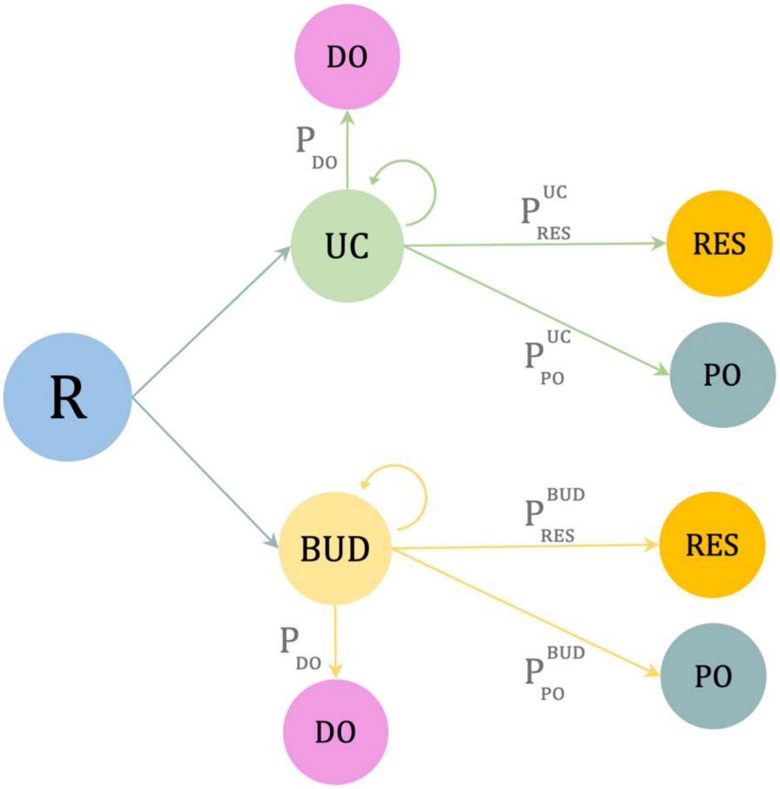


*Illustration 1*

*Patients are recruited (R) to either the BUD or UC arms and may, during each day of the virtual trial either remain recruited or have symptom resolution (RES), reach primary outcome (PO) or withdraw from the study (DO), according to probabilities, as shown.*

In particular, the ratio of $P^{UC}_{PO}$ to $P^{BUD}_{PO}$ represents a treatment effect, the purported reduction in the odds (on a daily basis) of reaching primary outcome that can be attributed to the effect of the budesonide treatment. We parametrised our model for the same number of patients recruited to both arms as in STOIC and worked backwards to estimate the maximum-likelihood daily probabilities for each of the transitions such that the mean virtual outcomes for the UC arm are the same, on average, as our findings (computer code available on request). We then studied the relationship between the ratio of $P^{UC}_{PO}$ to $P^{BUD}_{PO}$ and that of the ratios of patients reaching primary outcome during the virtual trial for the two arms. The results are illustrated in the main paper, Figure 5, with the grey envelope around the yellow curve (mean), representing the 95% confidence interval of outcomes. They indicate that in order to reach an average ratio of 10:1, a daily reduction in the odds of reaching primary outcome is approximately 3000%, with a minimum value (at 95% confidence) of approximately 400%, confirming that a very large daily treatment effect can be attributed to the use of budesonide inhalers for COVID-19 patients.

***Community detection algorithms***: To further elucidate the clinical trajectories of patients in the two arms of the STOIC study, we used community detection methods to interpret our finding as complex networks. Specifically, we treat each patient as a node in a network. The edges between nodes are averages of correlations between their time series in respect of a) highest daily temperature, b) lowest daily temperature, c) highest oxygen saturations, d) lowest oxygen saturations and e) heart rate. We weighted each of these time series correlations equally and only used data from days 1-14 because after this point, the data entries were sparser and yielded poor-quality correlation values.

In order to detect “communities” of patients that behave similarly to each other and differently to others, if these exist, we computed, over the resulting network, whose edges between two nodes *i* and *j* are denoted by $A_{ij}$, the maximum of a function of the type

$$Q(\vec{\sigma}) = \frac{1}{A_{\text{tot}}} \sum_{i,j} [A_{ij} - \langle A_{ij} \rangle] \delta(\sigma_i, \sigma_j)$$

Where $Q$ is the network modularity and $\delta$ indicates module membership, i.e. it is equal to 1 is two nodes belong to the same module. $A_{tot}$ is the sum over all the edge strengths computed as described above. This approach reveals a complex structure within our patient network, comprised of 4 modules, corresponding broadly to recovery trajectories (poor or effective) and each dominated by patients from either the BUD or UC arms (illustration 2).

This analysis is unique in that it looks at patient parameters over time in context, rather than at each in isolation. The results confirm qualitatively that the patients in the BUD arm recovered better in the totality of their measured parameters than did those in the UC arm, despite the small scale of our study. Isolated nodes indicate patients with sparsely recorded data or that dropped out and so did not fit in any of the communities

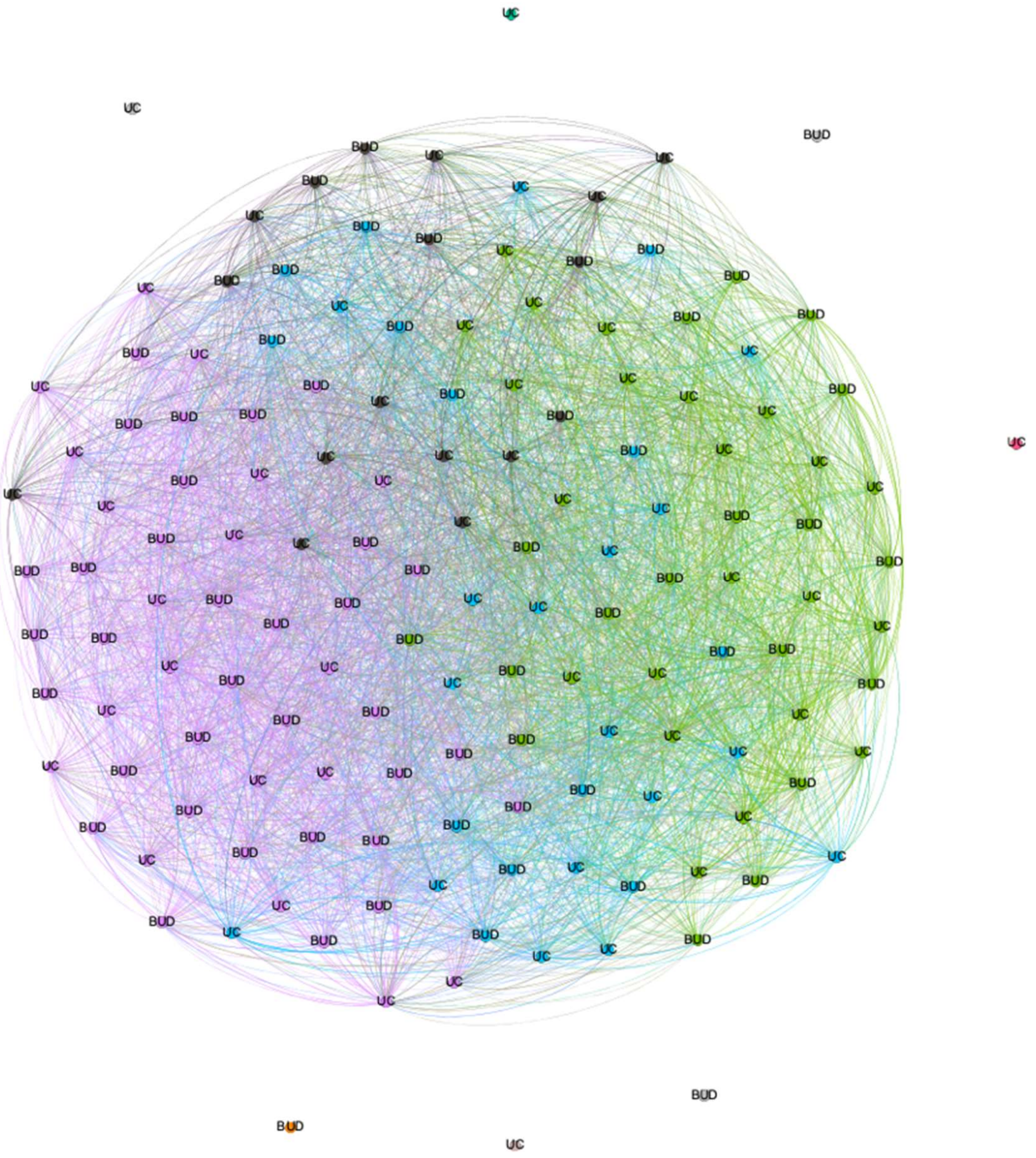
UC
BUD

**Supplementary Table 1**. Demographics and clinical characteristics of study participants in the intention to treat population

| Characteristic | Budesonide (n = 73) | Usual care (n = 73)[€] |
|---|---|---|
| Age, years | 44 (19-71) | 45 (19-79) |
| Female sex, no. (%) | 41 (56%) | 43 (61%) |
| White ethnicity, no. (%) | 67 (92%) | 66 (93%) |
| Body mass index, kg/m$^2$ | 26 (13 – 39) | 26 (18-42) |
| Number of co-morbidities, no. [¥] | 1 (range 0 - 5) | 1 (range 0 - 5) |
| Duration of symptoms prior to randomisation, days[¥] | 3 (Range 1 - 7) | 3 (range 0- 7) |
| Evidence of COVID positive status, no. (%) | 69 (96) | 68 (93) |
| Presence of symptoms at baseline, no. (%) | | |
| Cough | 58 (80%) | 49 (70%) |
| Fever | 52 (71%) | 45 (63%) |
| Headache | 41 (56%) | 38 (54%) |
| Fatigue | 33 (45%) | 24 (34%) |
| Loss of sense of smell/taste | 25 (34%) | 30 (42%) |
| Gastrointestinal symptoms | 12 (16%) | 12 (17%) |
| Breathlessness | 11 (15%) | 11 (16%) |
| Myalgia | 7 (10%) | 10 (14%) |
| Nasal symptoms | 3 (4%) | 5 (7%) |
| Sore throat | 0 (0%) | 2 (3%) |
| Chest pain/tightness | 4 (5%) | 1 (1%) |
| Other | 7 (10%) | 8 (11%) |
| FLUPRO score[*] | 0.818 (0.485) | 0.808 (0.443) |
| CCQ score[*] | 0.743 (0.441) | 0.668 (0.412) |
| Highest temperature recorded[*¥] in degrees centigrade | 36.6 (35.2-39.0) | 36.6 (35.5-38.3) |
| Lowest Oxygenation recorded[*¥] as % saturation | 96 (84%-99%) | 96 (90%-99%) |

Data presented as mean (SD) or mean (range) unless otherwise stated; [*]at randomisation [¥]Median (range); FLUPRO InFLUenza Patient Reported Outcome questionnaire; CCQ Common Cold Questionnaire; [€] 2 participants withdrew from study after study randomisation and only gender, age, and COVID-19 infection status was collected.

**Supplementary Table 2:** Demographics and clinical characteristics of study participants with a primary outcome compared to participants with symptom resolution in the per-protocol population

| **Characteristic** | **Primary outcome (n = 10)** | **Achieved symptom resolution (n = 129)** | **P value** |
|---|---|---|---|
| Age, years | 45 (19-79) | 45 (24-57) | 0.895 |
| Female sex, no. (%) | 9 (82%) | 71 (55%) | 0.085 |
| White ethnicity, no. (%) | 11 (100%) | 118 (92%) | 0.313 |
| Body mass index, kg/m$^2$ | 27 (18-42) | 26 (19-39) | 0.703 |
| Number of co-morbidities, no. ¥ | 1 (0-4) | 1 (0-4) | 0.904 |
| Duration of symptoms prior to randomisation, days¥ | 3 (2-7) | 3 (2-6) | 0.555 |
| Evidence of COVID positive status, no. (%) | 9 (90) | 120 (93) | 0.538 |
| Presence of symptoms at baseline, no. (%) | | | |
| Cough | 8 (73%) | 96 (74%) | 0.902 |
| Fever | 8 (73%) | 86 (67%) | 0.681 |
| Headache | 8 (73%) | 70 (54%) | 0.237 |
| Fatigue | 3 (27%) | 52 (40%) | 0.395 |
| Loss of sense of smell/taste | 4 (36%) | 51 (40%) | 0.836 |
| Gastrointestinal symptoms | 2 (18%) | 22 (17%) | 0.924 |
| Breathlessness | 3 (27%) | 19 (15%) | 0.273 |
| Myalgia | 2 (18%) | 14 (11%) | 0.463 |
| Nasal symptoms | 1 (9%) | 7 (5%) | 0.615 |
| Sore throat | 1 (9%) | 1 (1%) | 0.026 |
| Chest pain/tightness | 0 (0%) | 5 (4%) | 0.506 |
| Other | 2 (18%) | 13 (10%) | 0.404 |
| FLUPRO score* | 0.933 (SD: 0.333) | 0.806 (SD: 0.472) | 0.284 |
| CCQ score* | 0.800 (SD: 0.305) | 0.700 (SD: 0.437) | 0.354 |
| Highest temperature recorded*¥ in degrees centigrade | 36.6 (35.2 – 39.0) | 36.7 (35.8 to 37.2) | 0.870 |
| Lowest Oxygenation recorded*¥ as % saturation | 96 (84-99) | 96 (93-99) | 0.803 |

Data presented as mean (SD) or mean (range) unless otherwise stated; *at randomisation ¥Median (IQR); FLUPRO InFLUenza Patient Reported Outcome questionnaire; CCQ Common Cold Questionnaire

**Supplementary Table 3.** Delta mean change in FLUPro® symptoms between days 0 and 14 for the individual domains in the BUD and UC study arms.

| FLUPRO domain | Budesonide | Usual care | P value* |
|---|---|---|---|
| Systemic | -0.94 | -0.80 | 0.034 |
| Nose | -0.72 | -0.56 | 0.093 |
| Chest/Respiratory | -0.48 | -0.37 | 0.165 |
| Eyes | -0.28 | -0.23 | 0.325 |
| Throat | -0.61 | -0.57 | 0.542 |
| Gastrointestinal | -0.30 | -0.30 | 0.973 |

*from the ANCOVA model

**Supplementary Figure 1. Sensitivity analysis for time to clinical recovery in patients with confirmed SARS-CoV-2 infection**

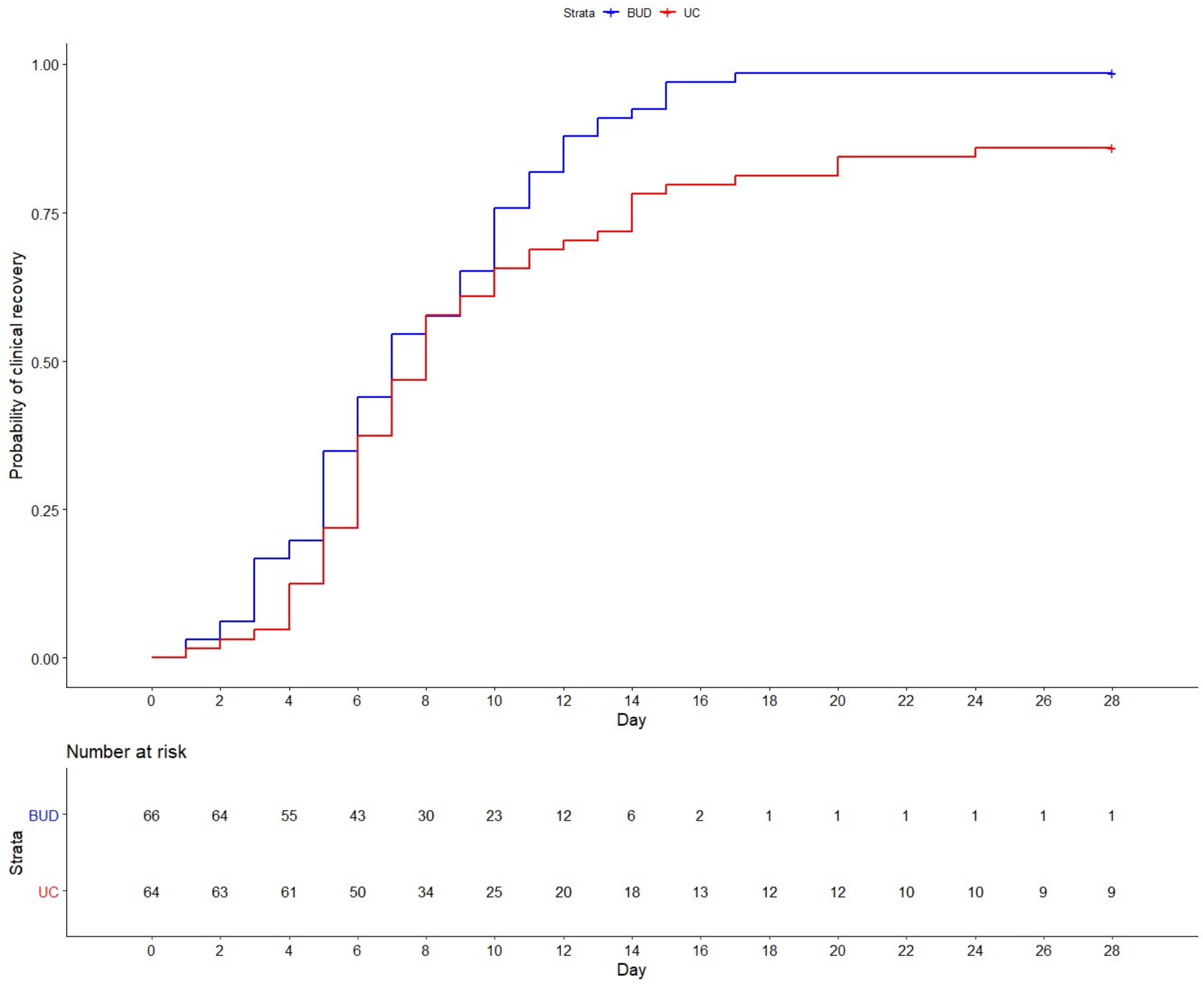

**Supplementary Figure 2.** *Daily peak temperature in BUD and UC participants. Trends indicate that daily highest temperature fell more rapidly in the BUD (-0.113 degrees Celsius per day) than the UC (-0.096 degrees Celsius per day) arm.*

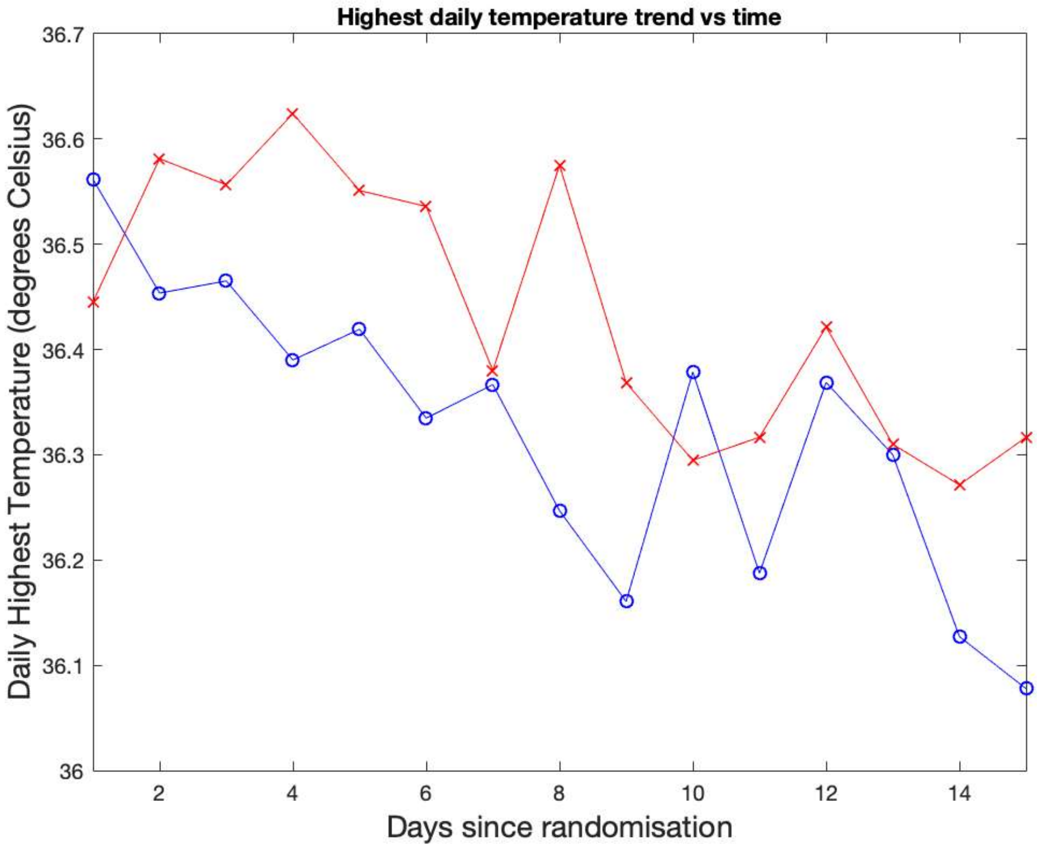

### Supplementary Figure 3. Time to symptom resolution as measured by the FLUPro®

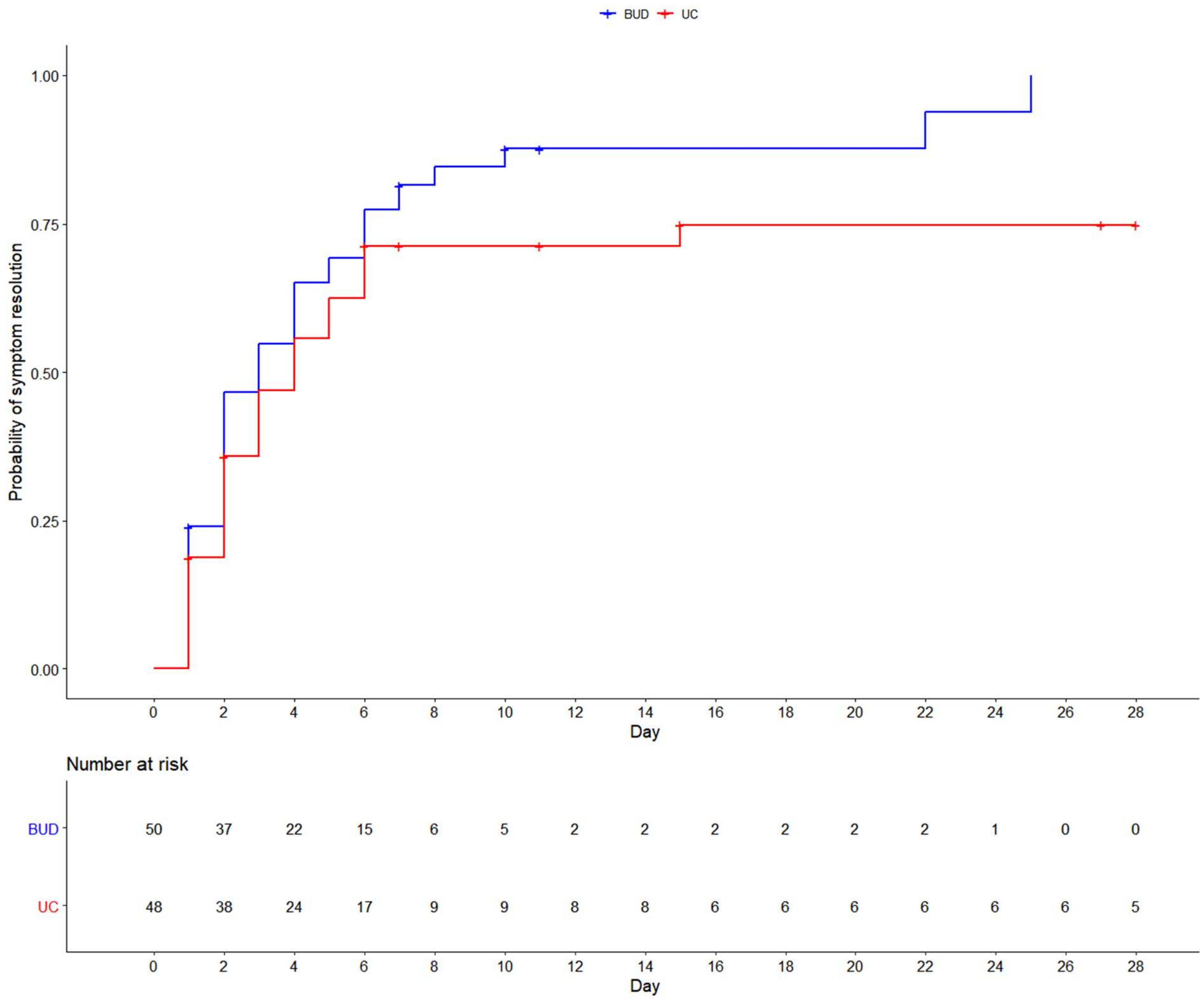

**Supplementary Figure 4. Daily mean Common Cold Questionnaire score for BUD and UC arm over 14 days**

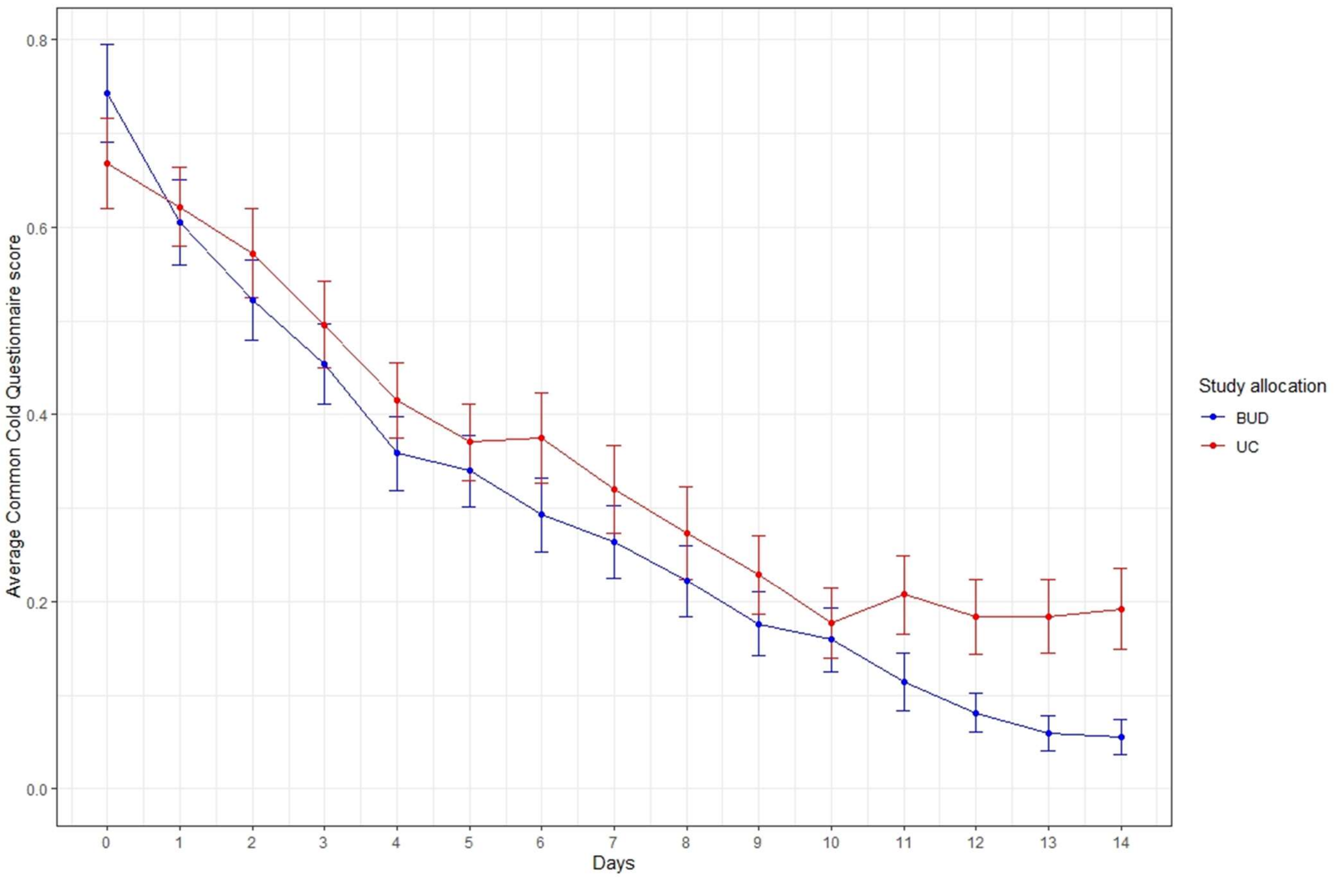

**Supplementary Figure 5.** Violin plots presenting Cycle Threshold (CT) over 3 study visits (day 0, 7 and 14) in the BUD and UC arm. Solid line represents median, dashed lines represent upper and lower interquartile. BUD = budesonide; UC = usual care. Lower limit of detection set at CT 40, CT values above 40 indicate undetectable virus

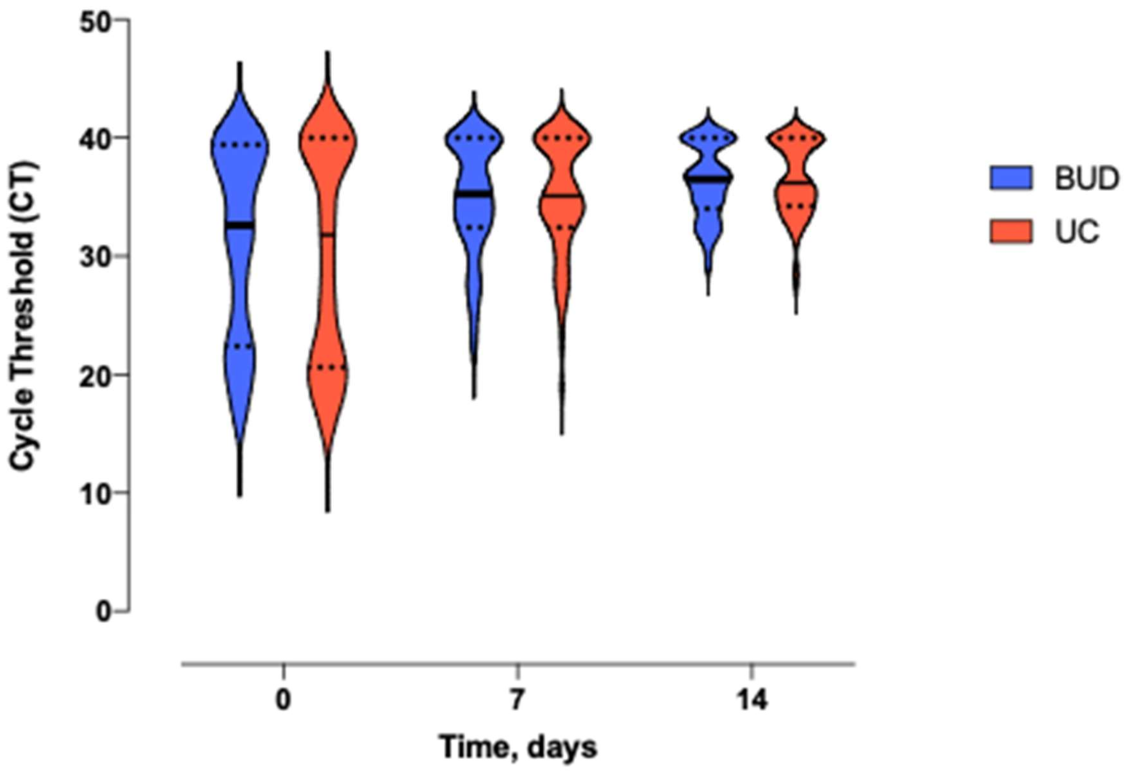
